# Biomakers for acute appendicitis: a two-sample Mendelian randomization study

**DOI:** 10.1101/2025.03.26.25324673

**Authors:** Ke Zhang, Hongwen OuYang

## Abstract

**Aim:** The causal link between acute appendicitis (AA) and a specific biomarker that can serve as an early diagnostic indicator remains unclear. The aim of this study was to evaluate potential causal effects of AA on various biomarkers using Mendelian randomization.

**Methods:** We performed a two-sample Mendelian randomization (MR) analyses using summary-level genome-wide association study (GWAS) data to explore the acute appendicitis on potential causal effects of 26 different biomarkers. A systematic evaluation and meta-analysis were performed for the positive biomarkers to assess the diagnostic accuracy for AA.

**Results:** Statistically significant association between the occurrence of AA and elevated levels of fibrinogen (FIB) (OR 1.1462, 95% confidence interval [CI] 1.0120 to 1.2982, P = 0.0318) and platelet count (PLT) (OR 1.0198, 95% confidence interval [CI] 1.0035 to 1.0363, P = 0.0169). The p-values of MR-Egger intercept analysis and Cochran’s Q test were both above 0.05. SROC curve of FIB and PLT in diagnosing AA were 0.75 [0.71-0.79] and 0.66 [0.61-0.70], respectively.

**Conclusion:** FIB and PLT have the potential to serve as early diagnostic biomarkers for acute appendicitis. However, further studies are required to determine the cut-off value and accuracy of their combined use in diagnosing acute appendicitis.

## Introduction

Acute appendicitis (AA) is a prevalent acute abdominal disease that can lead to serious complications such as bacterial peritonitis, sepsis, small intestinal obstruction, and abdominal abscess, among others [1]. The lifetime known risk of AA is 8.6% in men and 7.6% in women, with an annual incidence rate of 38.9% [2]. Accurate diagnosis and prompt treatment are crucial to improve overall outcomes for AA patients. The diagnosis of AA relies on a combination of anamnesis, physical examination, basic laboratory tests (including leukocyte count, neutrophil count, C-reactive protein [CRP], and procalcitonin [PCT]), as well as imaging tests such as abdominal ultrasound or abdominal CT scan [3–4]. However, despite the presence of well-established classical symptoms and clinical findings, early identification of acute appendicitis remains challenging and continues to be an area of difficulty [5].

Numerous researchers are currently engaged in the quest to develop a reliable tool or biomarker capable of accurately predicting the diagnosis of AA. Biomarkers such as fibrinogen, platelet count, mean platelet volume (MPV), interleukin-6 (IL-6), pentraxin-3 (PTX-3), and bilirubin have been investigated for their predictive potential and have recently demonstrated promising diagnostic accuracy and reliability, suggesting their relevance in the diagnosis of AA [3, 6–7]. However, it is important to note that the sensitivity and specificity of these biomarkers for early diagnosis of AA vary considerably across different centers, and their evaluation has largely been based on phenomenological descriptions. The causal link between AA and a specific biomarker that can serve as an early diagnostic indicator remains unclear. Further research is needed to confirm this and provide more evidence.

Mendelian randomization (MR) studies use genetic variations to assess causal relationship between the exposure and outcome. Genetic variants are allocated randomly at conception and confounding bias can be reduced by MR compared with conventional observational analyses. Single-nucleotide polymorphisms (SNPs) identified by genome-wide association studies (GWASs) were utilized as instrumental variables (IVs) [8]. If AA has causal effect on a specific biomaker, its genetically predicted levels should be as as a potential biomarker for early diagnosis of the disease.

The aim of this study was to evaluate potential causal effects of AA on various biomarkers using Mendelian randomization. We analyzed associations between AA and 26 different biomarkers by leveraging GWAS and FINNGEN data. Additionally, the accuracy of positive biomarkers in diagnosing AA was evaluated through a systematic review and meta-analysis.

## Methods

### Study Design and data sources

This study adheres to the Strengthening the Reporting of Observational Studies in Epidemiology using Mendelian Randomization (STROBE-MR) guidelines (additional checklist) [9–10]. The overall study design is presented in Figure 1. Two-sample Mendelian randomization (MR) analyses were conducted using summary-level genome-wide association study (GWAS) data. Since all data used in this study was publicly available, ethical approval was not required. Three key assumptions for instrumental variables were made: SNPs must be associated with the exposure, independent of confounding factors, and only affect the outcome through their influence on the exposure. A systematic search was performed to retrieve GWAS data from participants of European ancestry (up until May 2024) in the GWAS Catalog, UK Biobank, and FinnGen. The dataset of AA was used for the exposure variable, while 26 datasets of different biomarkers were included for the outcome variables. Only the largest GWAS summary statistics were included in the analysis.

**Figure 1.**
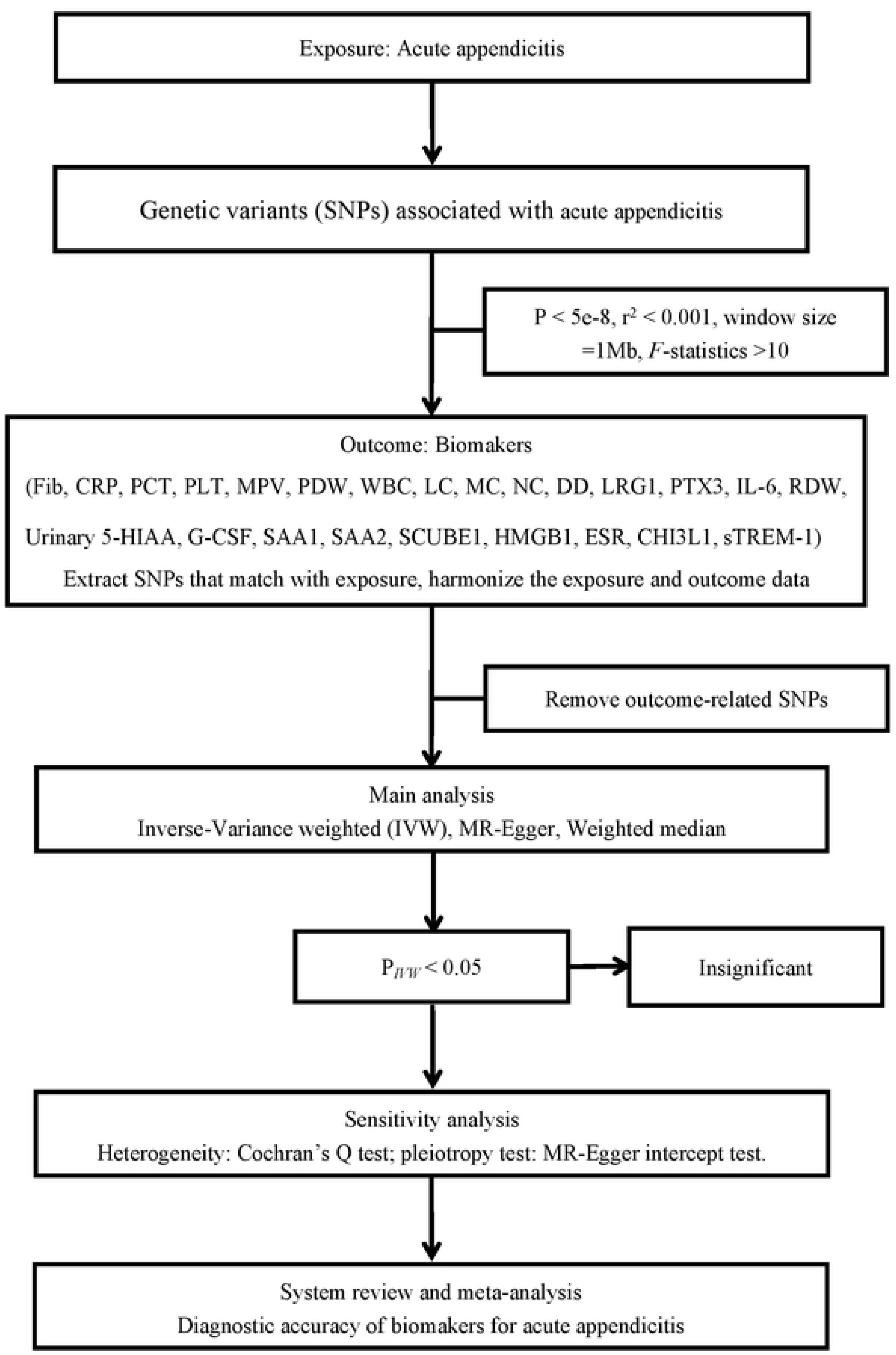
Overall design of the analysis framework in this study. Abbreviation: Fib, Fibrinogcn; CRP, C-reactive protein; PCT, Procalcitonin; PLT, Platelet count; MPV, Mean platelet volume; POW, Platelet distribution width; WBC, White blood cell count; LC, Lymphocyte count; MC, Monocytc count; NC, Ncutrophil count; DD, D-Dimcr; LRGI, Leucine-Rich Alpha-2-Glycoprotein; PTX3, Pentraxin-3; IL-6, lnterleukin-6 levels; ROW, Red Cell Distribution Width; Urinary 5-HIAA, Urinary 5-hydroxyindole acetic acid; G-CSF, Granulocyte colony stimulating factor; SAAI, Scrum amyloid A-I protein; SAA2, Scrum amyloid A-2 protein; SCUBEl, Signal peptide, CUB and EGF-like domain-containing protein I levels; HMGB I, High mobility group protein BI; ESR, Erythrocyte sedimentation rate; CHl3LI, Chitinasc 3-Likc I; sTREM-1, Soluble triggering receptor expressed on mycloid cells.

After identification and evaluation, summary-level dataset for AA was obtained from FinnGen (31,628 European cases, 378,082 controls) (https://www.finngen.fi/en/access_results). A total of 26 datasets for biomakers were retrieved from GWAS catalog: fibrinogen (n=10,708) [11], C-reactive protein (n=575,531) [12], procalcitonin (n=3,301) [13], platelet count (n=600,968) [14], mean platelet volume (n=694,866) [14], platelet distribution width (n=572,201) [14], White blood cell count (n=503,190) [14], lymphocyte count (n=509,277) [14], monocyte count (n=545,193) [14], neutrophil count (n=408,112) [15], D-Dimer (n=10,708) [11], leucine-rich alpha-2-glycoprotein (n=400) [16], pentraxin-3 (n=5,359) [17], interleukin-6 levels (n=14,743) [18], red cell distribution width (n=514,929) [14], bilirubin levels (n=56,577) [19], urinary 5-hydroxyindole acetic acid (n=4,910) [20], granulocyte colony stimulating factor (n=7,904) [21], serum amyloid A-1 protein (n=10,708) [11], serum amyloid A-2 protein (n=10,708) [11], Fetuin A (n=400) [16], Signal peptide, CUB and EGF-like domain-containing protein 1 levels (n=5,362) [17], High mobility group protein B1 (n=5,340) [17], erythrocyte sedimentation rate (n=10,541) [19], Chitinase 3-Like 1 (n=5,368) [17], Soluble triggering receptor expressed on myeloid cells (n=5,367) [17]. Full summary statistics for the outcomes are available for download from the GWAS catalog website (https://www.ebi.ac.uk/gwas/downloads/summary-statistics). Overview of GWAS used was shown in Supplement table 1.

### Mendelian Randomization

Independent genome-wide significant SNPs of AA were used as genetic instruments. Included instrumental variables should be strongly associated with the exposures (p ≤5×10^−8^) and SNPs with an F statistic <10 were excluded [22]. Linkage disequilibrium (LD) clumping was performed with r^2^<0.001 and window size of 10,000 kb [23]. The strength of each genetic instrument was assessed through the F statistic, calculated as F=R^2^ (N-2)/(1-R^2^), where R^2^ denotes the proportion of variance explained by the genetic instrument and N represents the effective sample size of the GWAS for the SNP-micronutrient association [22]. The R^2^ value was determined using the formula 2×MAF (1-MAF) beta^2^, where beta signifies the effect estimate of the genetic variant on the exposure, measured in standard deviation (SD) units, and MAF indicates the minor allele frequency [22–24]. Then, we removed SNPs associated with diseases or risk factors potentially associated with each biomaker (http://www.phenoscanner.medschl.cam.ac.uk/). The remaining SNPs were used in the MR analysis. The inverse variance-weighted (IVW) method was the main method for MR analysis and and the weighted median and MR-Egger methods were conducted to improve the IVW model-based estimation [24]. P<0.05 was considered nominally significant. Wald ratios for all SNPs were calculated. All MR analyses were performed using the TwoSampleMR package (version) in R (version) [25].

### Pleiotropy and and Sensitivity Analysis

We conducted MR-Egger regression to investigate the potential presence of pleiotropic effects of the SNPs. The intercept term in MR-Egger regression served as a valuable indicator to determine whether directional horizontal pleiotropy was influencing the results of the MR analysis. We utilized both the IVW method and MR-Egger regression to identify heterogeneity, and the degree of heterogeneity was measured using the Cochran Q statistic. A significance threshold of P < 0.05 was applied to indicate significant heterogeneity. Moreover, to identify any potentially influential SNPs, we conducted a leave-one-out test, whereby the MR was re-run while excluding each SNP in turn.

### System review and meta-analysis

After conducting Mendelian randomization analysis, a systematic evaluation and meta-analysis were performed for the positive biomarkers. The main objective was to assess the diagnostic accuracy of these positive biomarkers in diagnosing AA. This systematic review and meta-analysis strictly adheres to the guidelines by the Preferred Reporting Items for Systematic Reviews and Meta-Analyses (PRISMA) and Cochrane Handbook for Systematic Reviews of Diagnostic Test Accuracy [26–27]. We conducted a comprehensive search of Pubmed, Embase, and the Cochrane Database of Systematic Reviews up until May 1, 2024. The search was restricted to English language publications and had no restrictions on publication period.

Studies were included in meta-analysis with numbers of true positive, false positive, false negative and true negative test results to construct the 2×2 contingency table. Diagnostic performance parameters included sensitivity, specificity, positive likelihood ratio (PLR), negative likelihood ratio (NLR), diagnostic odds ratio (DOR) and area under the receiver operating characteristic (ROC) curve (AUC). Case series or reports, abstracts, duplication and conference presentations were excluded. The following data were extracted: author, year of published, study design, country, test method, sample size, age at diagnosis, cut-of value, numbers of true positive, false positive, false negative, true negative test results.

Quality Assessment of Diagnostic Accuracy Studies 2 (QUADAS-2) tool was used to evaluate the quality of studies [28]. The authors examined four domains: patient selection, index test, reference standard, flow and timing. The risk of bias in each domain was categorized as low, high or unclear. “Unclear” indicated an insufficient information to evaluate risk of bias. We resolved discrepancy by consensus.

Numbers of true-positive (TP), true-negative (TN), false positive (FP), and false-negative (FN) results were extracted from the study or calculated from reported diagnostic estimates. Pooled data of sensitivity, specificity, positive likelihood ratio, negative likelihood ratio, and diagnostic odds ratio were calculated using the random-effects model. Summary receiver operator curves (SROC) plots were made to summarize the diagnostic value of each parameter. Results of meta-analysis were presented as forest plots with effect size estimate and 95% confidence interval (95% CIs). I^2^ statistic was used to assess heterogeneity. Publication bias was evaluated with Deek’s funnel plot asymmetry test. The significance level was 0.05. The data lacking the standard deviation, conversion of correlation coefficients between Pearson and Spearman would be calculated based on formulas from the Cochrane handbook throughout the course [29–30]. Statistical analyses were performed using STATA/SE, version 16.0 and Review Manager 5.4.

## Results

### Mendelian Randomization

The detailed randomization analysis results between AA and biomarkers, including numbers of SNPs, ORs (95% CI) and p values, are presented in Supplement table 2. Forest plot results of all IVW are shown in Figure 2. Through multiple Mendelian randomization analyses, we observed statistically significant association between the occurrence of AA and elevated levels of fibrinogen (FIB) (OR 1.1462, 95% confidence interval [CI] 1.0120 to 1.2982, P = 0.0318) and platelet count (PLT) (OR 1.0198, 95% confidence interval [CI] 1.0035 to 1.0363, P = 0.0169). AA in this study were not shown to have significant associations with other biomakers. Information of SNPs after clumping process for FIB and PLT were show in Supplement table 3 and 4. The Results of pleiotropy and sensitivity analyses were shown in Supplement table 5 (P > 0.05). The p-values of MR-Egger intercept analysis and Cochran’s Q test were both above 0.05. The leave one out analysis and funnel plot of FIB and PLT were presented in Supplement Figure 1-4.

**Figure 2.**
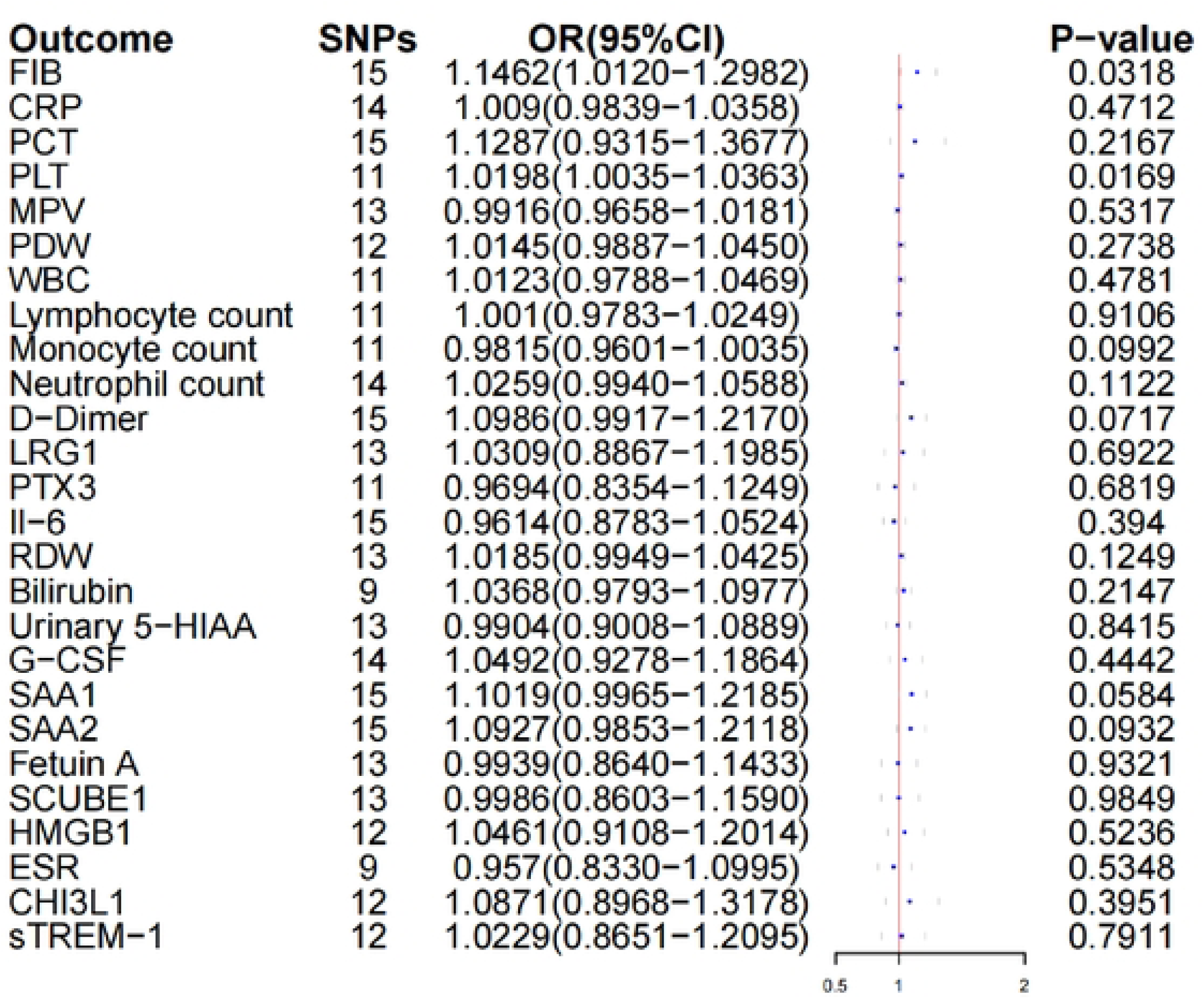
Forest plots showing causal estimates of acute appendicitis on biomakers.

### System review and meta-analysis

There were no studies available on the combined diagnostic use of FIB and PLT in AA. Only individual studies assessing the diagnostic utility of each biomarker in diagnosing AA were available. Eight studies included a total of 1324 patients were included in the review for FIB diagnosis of AA [31–38], and 7 studies included a total of 1531 patients were included in the review for PLT diagnosis of AA [38–44]. The PRISMA flow diagram were illustrated in Supplement figure 5 and 6. The characteristics of the included studies were summarized in Supplement table 6 and 7.

The cut-of value of FIB ranged from 245.5 to 520 ng/mL and PLT ranged from 188 to 263×10^9^/L. The sensitivity ranged from 43.90% to 88.14% and specifcity ranged from 47.06% to 92.80% for FIB, and the sensitivity ranged from 15% to 94.92% and specifcity ranged from 28.13% to 100% for PLT. Results of quality assessment by QUADAS-2 were shown in Supplement figure 7 and 8. The Pooled estimates of diagnostic performances of FIB and FIB for AA were calculated. Forest plots of sensitivity, specificity, positive likelihood ratio (PLR), negative likelihood ratio (NLR) and diagnostic odds ratio (DOR) were showed in Supplement figure 9 and 10. The sensitivity of FIB in the diagnosis of AA was 70% (95%CI, 0.59-0.81) [I^2^=91.40 (95%CI, 86.88-95.92)] and specificity was 69% (95%CI, 0.47-0.84) [I^2^=93.04 (95%CI, 89.61-96.48)]. The sensitivity of PLT in the diagnosis of AA was 56% (95%CI, 0.32-0.78) [I^2^=97.84 (95%CI, 97.04-98.64)] and specificity was 67% (95%CI, 0.40-0.86) [I^2^=94.28 (95%CI, 91.41-97.15)]. The results of sensitivity analysis were shown in Supplement table 8 and 9. Even after excluding any article, the heterogeneity was still high. SROC curve of FIB and PLT in diagnosing AA were illustrated in Supplement figure 11 and 12, which were 0.75 [0.71-0.79] and 0.66 [0.61-0.70].

## Discussion

The goal of modern surgical sciences is to achieve a balance between the complications of the disease and the complications of treatment. Previously, appendicitis was considered a surgical emergency, requiring immediate appendectomy to prevent perforation and other complications. However, the belief that the risk of perforation outweighs the negligible morbidity of negative laparotomy is now being disproven [45]. Therefore, the practice of rushing into surgery is no longer recommended, and observation and investigation are becoming more acceptable to ensure a more accurate diagnosis before surgical exploration. Several serum markers, imaging modalities, and clinical scoring systems have been introduced to reduce the rates of unnecessary appendectomies. However, their effectiveness in clinical practice has not been established yet. Inflammatory markers, such as white blood cell (WBC) count, C-reactive protein (CRP), procalcitonin (PCT), erythrocyte sedimentation rate (ESR), and neutrophil count, are commonly used tests in clinical practice. However, their elevation can be influenced by various conditions, making them nonspecific. Studies conducted on large populations have found that relying solely on these parameters results in unacceptably low specificity and sensitivity [46–47]. Mendelian randomization can help eliminate confounding factors and can be used to identify biomarkers that are causally associated with the occurrence of AA. These biomarkers can potentially serve as diagnostic markers for AA.

In this two-sample Mendelian randomization study, we found the occurrence of AA was causally associated with elevated levels of FIB and PLT. No causal effects of other factors involved in this study were found. Our study provided evidence on the potential early diagnostic biomarkers for acute appendicitis. We found that FIB and PLT individually exhibit a certain level of accuracy in diagnosing AA through our literature review. Unfortunately, it was found that there are currently no studies available on the accuracy of the combined use of both biomarkers in diagnosing AA.

FIB is an acute inflammatory mediator that is typically elevated in various acute inflammatory conditions, including acute appendicitis (AA) [48]. FIB not only plays a crucial role in hemostasis but also has pro-inflammatory functions. It can bind to receptors and activate immune cells involved in the inflammatory response [49]. FIB can act as a chemoattractant, recruiting polymorphonuclear cells (PMN) and fibroblasts, and it can activate mononuclear phagocytes. In conditions associated with infection, injury, and inflammation, FIB levels increase significantly. Studies have shown that FIB signaling during acute inflammation leads to the production of inflammatory cytokines, such as tumor necrosis factor-a and interleukin-1b [48–49]. These mediators have shown diagnostic utility in diagnosing AA. According to literature review, the sensitivity ranged from 43.90% to 88.14% and specifcity ranged from 47.06% to 92.80% for FIB diagnosis of AA. The final meta-analysis results reveal that the sensitivity was 70% and specificity was 69%.

Recent studies have highlighted the role of platelets in inflammation and their association with the pathophysiology of diseases involving inflammation [50]. Platelet count and derived indices, such as mean platelet volume (MPV) and platelet distribution width (PDW), can be measured using automated blood cell counters [51]. Although there have been reports in the literature about the diagnostic accuracy of MPV and PDW in diagnosing acute appendicitis (AA), this study, using a large sample Mendelian randomization, only found that PLT has potential in diagnosing AA. Several researchers found that platelet count was significantly higher in AA than in non-AA [39.42.44]. According to literature review in our study, the sensitivity ranged from 15% to 94.92% and specifcity ranged from 28.13% to 100% for PLT diagnosis of AA. The final meta-analysis results reveal that the sensitivity was 56% and specificity was 67%.

However, the heterogeneity among included studies was high, which may be attributed to differences in patient populations, study designs, and cut-off values of FIB and PLT. The difference in cut-of value plays an important role in the apparent heterogeneity. In the literature we included in the analysis, the cut-of value of FIB ranged from 245.5 to 520 ng/mL and PLT ranged from 188 to 263×10^9^/L. Although sensitivity analysis was conducted (Supplement table 8 and 9), the heterogeneity remained high, which indicated the need for further studies to validate the outcomes.

There are limitations in out study. Firstly, there may be other reliable biomarkers for diagnosing AA that are currently unknown, as the 26 biomarkers included in this study were only obtained through literature searches. Secondly, the sample size for some biomarkers, such as PTX3 and Fetuin A, was relatively small, with only 400 participants included in the dataset, requiring more large sample size accumulation for subsequent analysis. Thirdly, there is currently no research on using FIB in conjunction with PLT to diagnose AA, with only individual reports on the diagnosis of AA, and the sample sizes and cut-off values reported in the literature are not consistent. Fourthly, the heterogeneity of the meta-analysis results in this study is very high, and more large sample size multicenter studies are needed to verify the results of our study, especially for the accuracy of combined diagnosis of AA using both biomarkers.

## Conclusions

Fibrinogen and platelet count have the potential to serve as early diagnostic biomarkers for acute appendicitis. However, further studies are required to determine the cut-off value and accuracy of their combined use in diagnosing acute appendicitis.

## Data Availability

All relevant data are within the manuscript and its Supporting Information files

